# Non-Invasive Arterial Blood Pressure Waveform Generation in Critically Ill Patients: A Sensor-Based Deep Learning Approach

**DOI:** 10.64898/2026.04.28.26351954

**Authors:** Carl Harris, Bright Nnadi, Sampath Rapuri, John Rattray, Francesco Tenore, Ralph Etienne-Cummings, Robert D. Stevens

## Abstract

Continuous monitoring of Arterial Blood Pressure (ABP) in critically ill patients requires invasive arterial catheterization, which carries risks of thrombosis, vascular injury and infection. Here, we train and validate a computational model for continuous non-invasive ABP estimation in Intensive Care Unit (ICU) patients using a novel wearable sensor array. The sensor acquires continuous high frequency photoplethysmography (PPG) and electrocardiography (ECG) signals which are used as inputs in a deep learning algorithm for beat-to-beat reconstruction of ABP waveforms. We include 28 patients enrolled in four ICU units at Johns Hopkins Hospital, comprising 15,489 five-second ECG and PPG segments. A CNN/LSTM hybrid architecture achieved an *R*^2^ of 0.812 and a sample-level mean absolute error (MAE) of 4.94 ± 4.96 mmHg, with systolic and diastolic blood pressure MAEs of 6.38 ± 6.62 and 3.99 ± 4.53 mmHg, respectively. This performance closely approached an upper-bound model trained on contemporaneously acquired ground truth ECG and PPG signals (*R*^2^ = 0.824, *MAE* = 4.81 mmHg), indicating that the sensors retain most hemodynamically relevant information. Split-conformal prediction provided calibrated uncertainty intervals with coverage meeting nominal targets, offering a principled framework for bedside confidence assessment. These findings demonstrate the feasibility of accurate, continuous, non-invasive ABP waveform estimation from wearable biosignals in critically ill patients, establishing a foundation for reducing dependence on invasive arterial monitoring while preserving the waveform-level information essential for hemodynamic management.

## Introduction

Blood Pressure (BP) represents one of the most fundamental physiological parameters in clinical medicine, serving as a critical indicator of cardiovascular health and hemodynamic stability. Hypertension is the leading modifiable risk factor for cardiovascular disease and premature mortality worldwide, affecting over one billion adults globally and contributing to approximately 10 million preventable deaths annually^1,2^. In the United States alone, hypertension is associated with population-level healthcare expenditures exceeding $131 billion per year, representing a substantial economic burden on the healthcare system^3^. Despite widespread awareness and the availability of effective antihypertensive medications, less than a quarter of hypertensive individuals maintain adequate blood pressure control, with approximately 70% of patients requiring combination therapy with multiple agents to reach target levels^4^.

The clinical importance of precise blood pressure monitoring is particularly pronounced in the Intensive Care Unit (ICU), where both extremes of blood pressure carry significant consequences. Acute elevations in blood pressure can precipitate hypertensive emergencies with downstream consequences that include ischemic and hemorrhagic stroke, acute myocardial infarction, aortic dissection, and pulmonary edema, and death^5,6^. Conversely, hypotension in critically ill patients leads to inadequate tissue perfusion, resulting in organ dysfunction, shock, and increased mortality^7^. The dynamic nature of blood pressure in acutely ill patients, combined with the narrow therapeutic windows for vasoactive medications, necessitates continuous, beat-to-beat monitoring to guide clinical decision-making and optimize outcomes.

The gold standard for continuous blood pressure monitoring is invasive arterial catheterization, in which a cannula is inserted directly into a peripheral artery, enabling real-time pressure waveform acquisition with high temporal resolution^8^. Beyond providing systolic and diastolic values, Arterial Blood Pressure (ABP) waveforms contain rich morphological information that reflects cardiac function, vascular compliance, and overall hemodynamic status. Analysis of waveform characteristics such as the dicrotic notch, pulse pressure, and waveform contour can provide insights into stroke volume, cardiac output, and arterial stiffness^9^. Despite its clinical utility, invasive arterial monitoring introduces significant risks including infection, thrombosis with potential distal ischemia, bleeding, hematoma formation, and arterial damage^10,11^. In the United States, an estimated 80,000 catheter-related bloodstream infections occur annually, and while major complications are relatively rare (occurring in fewer than 1% of placements), the cumulative risk increases with prolonged catheterization^12^. These concerns motivate the development of non-invasive alternatives that could reduce or eliminate the need for arterial line placement while preserving the benefits of continuous waveform monitoring.

Recent advances in wearable sensing technologies and machine learning have opened new avenues for non-invasive estimation of blood pressure from peripheral biosignals. Photoplethysmography (PPG), which uses optical sensors to detect volumetric changes in blood in the microvasculature, has emerged as a particularly promising modality due to its ease of acquisition, low cost, and established presence in consumer devices^13^. The PPG waveform morphology reflects arterial pressure wave propagation and contains information that correlates with underlying circulatory parameters. When PPG is acquired simultaneously in two anatomical sites or with electrocardiography (ECG), BP can be computed using simple equations such as the pulse arrival time (PAT) and pulse transit time (PTT), which evaluate the arterial stiffness-pressure relationship^14,15^. These cardiovascular timing intervals form the physiological basis for many cuffless blood pressure estimation approaches. Alternative approaches, such as the Finapres system^16^, use a volume-clamp technique on the finger to provide continuous beat-to-beat blood pressure measurements at higher sampling rates. However, the Finapres requires a tethered connection to a bedside unit and necessitates frequent calibration against an upper-arm cuff, limiting its practicality for ambulatory or long-term monitoring^17^. These constraints underscore the appeal of fully wearable, calibration-free methods based on PPG and ECG.

The work presented here is a novel machine learning framework for non-invasive estimation of continuous ABP waveforms using PPG and ECG signals recorded from hospitalized patients with concurrent invasive arterial monitoring. We test the hypothesis that a deep learning model trained on PPG and ECG signals acquired from a novel wearable sensor array can accurately reconstruct continuous ABP waveforms in critically ill ICU patients. We further evaluate whether split-conformal prediction can provide calibrated, clinically interpretable uncertainty estimates for these predictions, and assess the model’s ability to detect hemodynamically significant blood pressure derangements.

Building upon the MOSAIC (MultimOdal Sensor Array for Integrated Computation) sensor architecture previously developed for ambulatory blood pressure estimation^18,19^, we extend this methodology to the ICU setting where ground-truth ABP waveforms enable rigorous model training and validation. Our approach employs a deep learning architecture optimized for waveform reconstruction, with careful attention to signal quality assessment and physiologically informed preprocessing. We evaluate model performance not only on standard regression metrics but also on clinically relevant classification tasks including detection of hypertensive and hypotensive episodes, which directly impact patient management decisions.

## Methods

### Recording paradigm

Data were collected in ICUs at Johns Hopkins Hospital (JHH) under a protocol approved by Johns Hopkins Medicine Institutional Review Board-approved protocol (IRB00384821). Patients or their legally authorized surrogates who provided written consent were enrolled in multiple critical care units including the Weinberg ICU (WICU), Surgical ICU (SICU), Cardiovascular Surgery ICU (CVSICU), Cardiac Care Unit (CCU). Enrollment required the patient or a legally authorized surrogate to provide informed consent. Eligible patients were adults aged 18 years or older who were admitted to one of these units with an arterial catheter already in place as part of standard clinical care. Patients were excluded if extensive injuries, disease, or wound dressings involving the chest or hands precluded appropriate sensor placement, or if they belonged to a vulnerable population (e.g., prisoners, wards of state, or assault victims).

The MOSAIC sensor array was applied to enrolled patients for continuous data collection. Devices were placed on the chest and one finger. The device placed on the chest captured ECG, while the sensor placed on the finger recorded PPG signals. Both sensors recorded accelerometry data as well, though this input signal was not included in our current analyses. Sensors wirelessly transmitted data via Bluetooth Low Energy (BLE) to a central computer for real-time aggregation and storage. Concurrent invasive arterial blood pressure (ABP) waveforms, serving as ground truth, were accessed from the JHH Sickbay clinical data repository and time-aligned with the wearable sensor recordings.

### Population of interest

The target population consisted of critically ill adults requiring invasive hemodynamic monitoring in the ICU. This setting was selected because the continuous, beat-to-beat ABP waveforms available from indwelling arterial catheters provide the high-fidelity, waveform-level ground truth necessary for training and rigorously validating a machine learning model for non-invasive ABP estimation. Critically ill patients represent a clinically high-stakes population in whom accurate, continuous blood pressure monitoring is essential: acute hypertension in this setting can precipitate stroke, myocardial infarction, and aortic dissection, while hypotension leads to organ hypoperfusion, acute kidney injury, and increased mortality. At the same time, this population encompasses a broad and heterogeneous range of hemodynamic states, including vasopressor-dependent shock, post-operative recovery, and neurological emergencies, making it an appropriate and challenging benchmark for evaluating model generalizability.

### Data Preprocessing

For each trial, continuous waveforms from the wearable sensor (ECG and PPG) and from the High-Frequency (HF) bedside monitor were first brought onto a common time base. The temporal overlap among the sensor ECG, sensor PPG, and HF recording was computed; trials with no valid overlap were excluded. Within the overlapping interval, a reference timeline was defined by the shorter of the two sensor streams (ECG or PPG), truncated to that duration so that every reference sample had valid sensor data (i.e., overlapping sensor ECG, PPG, and ground-truth ABP).

Because wearable and bedside signals can exhibit clock drift and subtle temporal misalignment, a second alignment step was applied to improve sample-level correspondence between sensor and ground-truth ECG. R-peaks were detected on both the sensor and ground-truth ECG using NeuroKit2^20^. A greedy matching algorithm then associated each ground-truth R-peak with the nearest unmatched sensor R-peak, accepting only pairs whose separation did not exceed a maximum distance set to 50% of the mean R-R interval estimated from the sensor peaks. From the resulting set of matched (sensor index, ground-truth index) pairs, a piecewise linear mapping from sensor sample indices to ground-truth sample indices was built, with boundary extrapolation at the start and end of the recording. This mapping was used to compute, for each sensor sample index, the corresponding ground-truth index; ground-truth ECG, ABP, and PPG were then interpolated (linear interpolation) onto these aligned indices and written as the final time-aligned waveforms. All processed files were passed through this pipeline to produce synced datasets used for segmentation and modeling.

To restrict analysis to periods where the wearable ECG agreed well with the reference, the synced data were segmented into non-overlapping 5-minute blocks. For each block, the Pearson correlation coefficient between the sensor ECG and the ground-truth ECG was computed (after excluding samples with missing values). Blocks whose correlation fell below a fixed threshold of 0.3 were discarded; blocks meeting or exceeding this threshold were retained for downstream segmentation and model training or evaluation (see **Figure S1**).

Segments were excluded if the ground-truth ABP waveform contained any sample outside the physiologic range of 20-300 mmHg. This criterion removed artifactual or non-physiologic ABP segments from the analysis set. Per-segment signal quality indices (SQI) were computed for both ECG and PPG to retain only windows with sufficient quality. For ECG, the segment was cleaned with NeuroKit2, R-peaks were detected^21^, and the average-QRS quality metric was evaluated; the mean of this quality time series over the segment was taken as the ECG SQI. For PPG, the segment was cleaned, peaks were detected^22^, and the template-match quality metric was computed; the mean over the segment was taken as the PPG SQI. A segment was retained only if both ECG SQI and PPG SQI were greater than or equal to a threshold of 0.6. Segments for which SQI computation failed (e.g., insufficient peaks) or which did not meet the threshold for either channel were excluded. A detailed flowchart of sample selection criteria is presented in **Figure 1A**.

**Figure 1:**
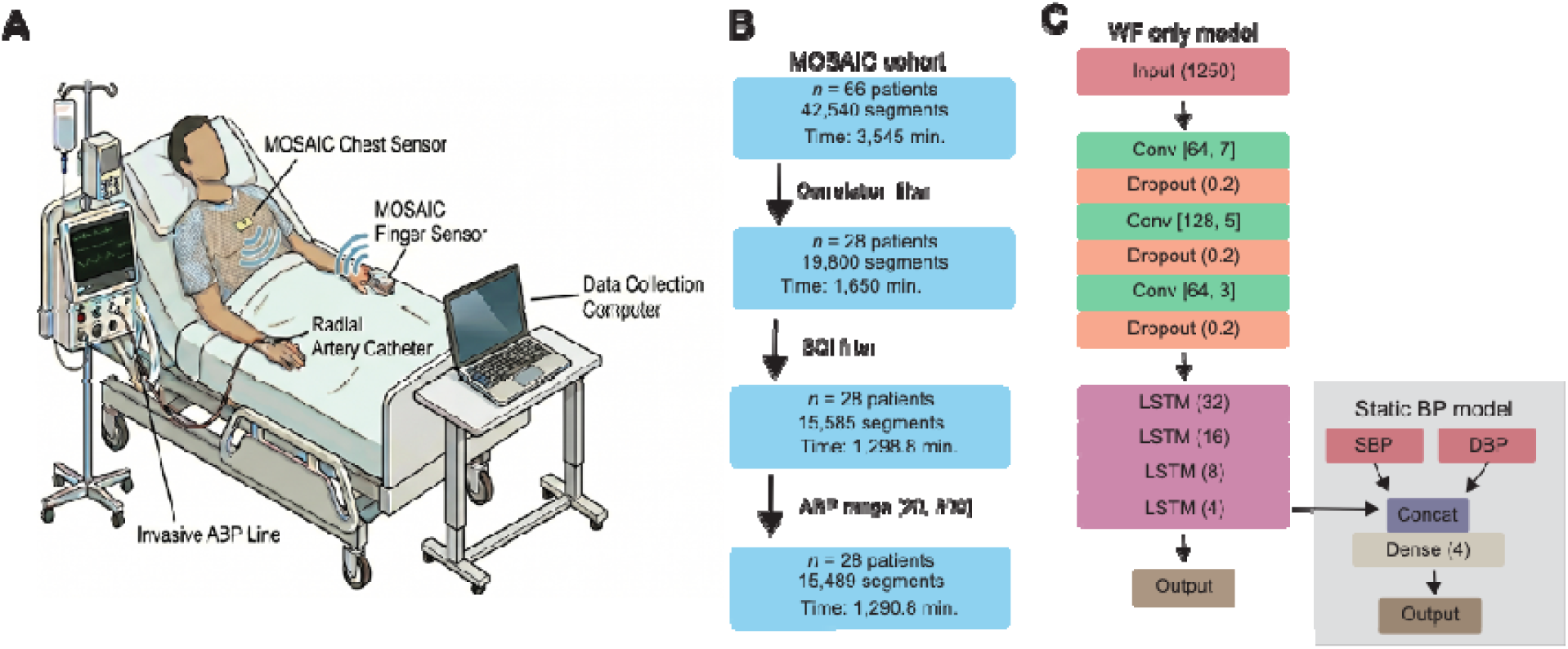
Study sample selection and model architecture. **(A)** Schematic of the MOSAIC sensor setup, which consists of two sensors (chest and finger) that transmit to a data collection laptop, and an arterial line measurement that records groundtruth values. **(B)** Overview of the sample selection pipeline. Sixty-six patients with indwelling arterial catheters were enrolled across four ICU units at JHH and recorded using the MOSAIC wearable sensor array. Continuous ECG and PPG signals from the wearable devices were time-aligned with invasive arterial blood pressure (ABP) waveforms from the bedside monitor via R-peak–based piecewise linear registration. Recordings were divided into non-overlapping 5-minute blocks; blocks with sensor-to-reference ECG correlation below 0.3 were discarded. Within retained blocks, segments with ABP values outside the physiologic range (20–300 mmHg) or with ECG or PPG SQIs below 0.6 were excluded, yielding the final analysis cohort of 28 patients. **(C)** Model architecture. Each 5-second segment (1,250 samples at 250 Hz) comprising two input channels (ECG and PPG) was passed through four stacked bidirectional LSTM layers (hidden sizes 32, 16, 8, and 4). Block-level systolic and diastolic blood pressure values were encoded through a dense layer (4 units, ReLU activation) and broadcast across all time steps. The concatenated sequential and static features (8 dimensions per time step) were projected through a final dense layer to produce the predicted ABP waveform at each time point.

The synced, filtered data were segmented in two stages. First, the continuous record was divided into non-overlapping 5-minute blocks. Within each retained block, non-overlapping 5-second windows were extracted, yielding segments of 1,250 samples per window at a sampling rate of 250 Hz. Each 5-second segment thus contained two input channels (ECG and PPG, either from the sensor or from the ground-truth waveforms, depending on the experiment) and the corresponding ground-truth ABP waveform as the regression target.

### Static model

In addition we introduce a secondary model (see **Supplementary Tables**) that include two static covariates, systolic and diastolic blood pressure (SBP and DBP), that were associated with the beginning every segment belonging to a given 20-minute block. The idea here was to simulate cuffed blood pressure measurements, which could be sampled every 20 minutes and serve as a calibration point for our algorithm. These were derived from the first complete cardiac cycle within that block: R-peaks were detected on the ground-truth ECG, the first inter-peak interval was used to define the cycle, and SBP and DBP were taken as the maximum and minimum of the ground-truth ABP over that cycle.

### Model architecture and training

We used a hybrid CNN-bidirectional LSTM model for sequence-to-sequence prediction of the ABP waveform from concurrent ECG and PPG. The sequential input is a 5-second window of two channels (ECG and PPG) at 250 Hz (1,250 time steps × 2 features). The sequence is first passed through three one-dimensional convolutional layers (64, 128, and 64 filters with kernel sizes 7, 5, and 3, ReLU activation, same padding, and dropout 0.2 after each), then through four stacked bidirectional LSTM layers and hidden sizes 32, 16, 8, and 4 (dropout and recurrent dropout 0.2 on the first three, dropout 0.1 on the last). The primary model has 105,209 trainable parameters.

All sequential inputs, as well as the target ABP, were standardized (zero mean, unit variance) using the training set only; the same scaling parameters were applied to validation and test data. The loss function was a mean squared error (MSE). Training was performed with the Adam optimizer (learning rate 0.001), with early stopping on validation loss (patience 5 epochs, restoring best weights) and reduction of learning rate on plateau (factor 0.5, patience 3 epochs, minimum learning rate 1x 10^-6^). Batch size was 64. The number of epochs was determined by early stopping.

### Model evaluation

Evaluation was performed on a held-out test set of segments. All metrics were computed after inverse-transforming predictions and ground truth to the original ABP scale (mmHg). At the sample level, we report the coefficient of determination (*R*^2^), mean absolute error (MAE), SBP, and DBP, between the flattened predicted and true ABP sequences. A simple baseline was defined by predicting the training-set mean ABP at every time point for every segment; its MAE was reported for comparison. We further defined a number of comparison models (e.g., dummy regressor, pure LSTMs of varying sizes, and a previous model which integrates PPG and ECG to predict ABP^23^); additional details are described in the **Supplemental Methods**.

Primary metrics were MAE (mmHg), and *R*^2^ for the continuous ABP waveform, as well as MAE for SBP and DBP. Secondary metrics included AU-ROC for binary classification of hypertension and hypotension at the segment level, based on SBP and DBP.

### Uncertainty quantification

To provide prediction intervals with guaranteed coverage properties and to quantify uncertainty in a way that is useful at the bedside, we applied split-conformal prediction with a conditionally varying scale. The data were split into training, calibration, and test sets: after holding out 20% of segments for testing, the remainder was further split so that a dedicated calibration set (e.g., 20% of the training portion) was used only for conformal calibration and never for model fitting. On the calibration set, the model produced point predictions for the full ABP waveform; at each (segment, time) pair we computed the absolute residual *R* = |*y* - *ŷ*|. To allow interval width to depend on the predicted value, so that intervals are wider where the model tends to err more, we estimated a conditional scale *σ*(*ŷ*) by binning calibration predicted values into 40 percentile-based bins and, within each bin, taking the median absolute residual. Bins with fewer than 10 samples used the global median residual instead, and the scale was lower-bounded by a small constant to avoid division by zero. The nonconformity score was defined as *E = R*/*σ*(*ŷ*), i.e., the residual normalized by this conditional scale. The (1 - *α*) quantile q of the set of calibration nonconformity scores was then computed (with α the target miscoverage rate, e.g., 0.1 for 90% coverage), using the standard finite-sample formula so that the resulting intervals achieve at least (1 - *α*) coverage under exchangeability. At test time, for each predicted value *ŷ*(*t*) we assigned the same *σ*(*ŷ*) via the precomputed bin edges and bin scales; the prediction interval at that time point was [*ŷ*(*t*) -*q* ·*σ*(*ŷ*), *ŷ*(*t*)+ *q· σ*(*ŷ*). Thus, the half-width *q* ·*σ*(*ŷ*) varies with the prediction: it is larger when the calibration residuals in that predicted-value bin were larger, providing a data-driven measure of uncertainty that adapts to BP level.

We evaluated empirical coverage at several levels. Pointwise coverage was the proportion of all test-set ABP samples (across segments and time) that fell within their respective intervals. Segment-level coverage was computed for systolic and diastolic pressure: for each test segment, SBP and DBP were taken as the maximum and minimum of the ground-truth ABP over the 5-second window, and we checked whether each lay within the interval derived from the corresponding predicted SBP or DBP (using the same *q* and the *σ*(*ŷ*) evaluated at the predicted SBP/DBP). These empirical coverage rates were compared to the theoretical target 1 - *α* to assess calibration. In addition, we used the conformal interval width as a proxy for uncertainty: for each segment, the widths of the SBP and DBP intervals were classified into tertiles (narrow, moderate, wide) based on the distribution of widths in the test set, so that clinicians could distinguish segments where the model was relatively confident (narrow intervals) from those where uncertainty was high (wide intervals). This combination of distribution-free coverage guarantees (under exchangeability) and conditionally varying width provides both rigorous uncertainty quantification and interpretable, value-dependent intervals for downstream decision support.

Results of the conformal calibration, including the estimated quantile *q*, the range of *σ*(*ŷ*) on the calibration set, and the comparison of empirical versus theoretical coverage for pointwise, SBP, DBP, and joint segment-level coverage, are reported in the **Results**. Example test segments were plotted with the predicted ABP waveform and the conformal band (*ŷ* ± *q*· *σ*(*ŷ*)) shaded, so that interval width could be inspected over the cardiac cycle.

## Results

Of 66 patients enrolled in the MOSAIC cohort, 28 met all inclusion criteria and contributed analyzable data to the study. Demographic and clinical characteristics of the enrolled cohort are presented in **Table 1**. After application of the signal quality filters (ECG-ground truth correlation ≥ 0.3; ECG SQI ≥ 0.6; PPG SQI ≥ 0.6) and exclusion of segments with non-physiologic ABP values, the final dataset comprised 15,489 five-second segments available for model training and evaluation. The MOSAIC cohort consists of all patients enrolled in the study, while the “Analysis cohort” includes patients who matched the sample selection criteria described in the **Methods** section.

**Table 1.**
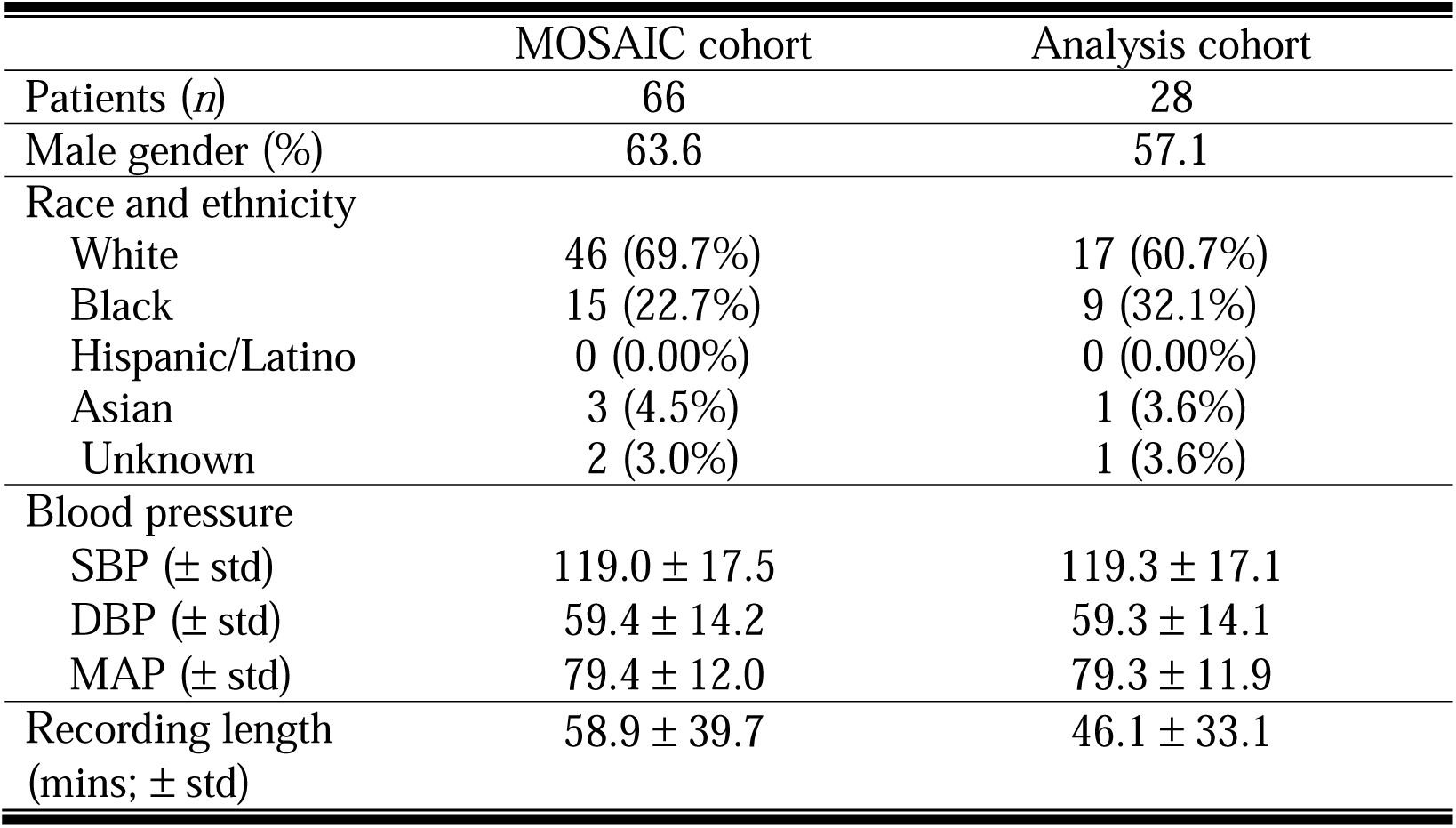
Population sample characteristics.

|  | MOSAIC cohort | Analysis cohort |
| --- | --- | --- |
| Patients (*n*) | 66 | 28 |
| Male gender (%) | 63.6 | 57.1 |
| Race and ethnicity |  |  |
| White | 46 (69.7%) | 17 (60.7%) |
| Black | 15 (22.7%) | 9 (32.1%) |
| Hispanic/Latino | 0 (0.00%) | 0 (0.00%) |
| Asian | 3 (4.5%) | 1 (3.6%) |
| Unknown | 2 (3.0%) | 1 (3.6%) |
| Blood pressure |  |  |
| SBP (± std) | 119.0 ± 17.5 | 119.3 ± 17.1 |
| DBP (± std) | 59.4 ± 14.2 | 59.3 ± 14.1 |
| MAP (± std) | 79.4 ± 12.0 | 79.3 ± 11.9 |
| Recording length<br>(mins; ± std) | 58.9 ± 39.7 | 46.1 ± 33.1 |

### Primary model performance

The CNN/LSTM hybrid (CNN/LSTM hybrid) achieved the best overall waveform reconstruction among all sensor-input models, with an *R*^2^ of 0.812 and a sample-level MAE of 4.94 ± 4.96 mmHg (**Table 2**). This represented a substantial improvement over the dummy (mean-prediction) baseline, which achieved an *R*^2^ of 0.001 and an MAE of 15.01 ± 5.03 mmHg, confirming that the model captured meaningful hemodynamic signal beyond trivial population-level statistics. The CNN/LSTM hybrid model also achieved SBP and DBP MAEs of 6.38 ± 6.62 mmHg and 3.90 ± 4.53 mmHg, respectively.

**Table 2.** Comparison of model architectures for arterial blood pressure waveform reconstruction. Performance metrics for all evaluated models on the held-out test set, including the coefficient of determination (R²), sample-level mean absolute error (MAE ± SD, mmHg), and segment-level SBP and DBP. Models are grouped by input source: ground-truth bedside monitor signals (GT) serving as an upper-bound reference, and wearable MOSAIC sensor signals for all remaining architectures. LSTM models are reported at three capacity levels: small (hidden sizes 32/16/8/4), medium (64/32/16/8), and large (128/64/32/16). The dummy regressor predicts the training-set mean ABP for all samples and serves as a naive baseline.

| Model | $R^2$ | MAE | SBP MAE | DBP MAE |
| --- | --- | --- | --- | --- |
| Dummy regressor | 0.001 | $15.01 \pm 5.03$ | $40.94 \pm 12.75$ | $20.06 \pm 8.26$ |
| GT | 0.824 | $4.81 \pm 4.53$ | $5.76 \pm 6.02$ | $3.55 \pm 4.98$ |
| LSTM (small) | 0.741 | $6.29 \pm 5.12$ | $7.72 \pm 7.76$ | $4.57 \pm 5.74$ |
| LSTM (med) | 0.778 | $5.60 \pm 5.05$ | $7.02 \pm 7.18$ | $3.88 \pm 5.57$ |
| LSTM (large) | <u>0.787</u> | <u>$5.34 \pm 5.11$</u> | <u>$6.51 \pm 7.10$</u> | <u>$3.78 \pm 5.60$</u> |
| Transformer | 0.769 | $5.90 \pm 6.38$ | $7.09 \pm 8.32$ | $3.84 \pm 6.82$ |
| CNN/LSTM hybrid | **0.812** | **$4.94 \pm 4.96$** | **$6.38 \pm 6.62$** | **$3.99 \pm 4.53$** |
| UNET | 0.507 | $9.15 \pm 5.16$ | $10.55 \pm 9.32$ | $4.44 \pm 5.81$ |
| Yen et al. (2022)<sup>23</sup> | 0.4152 | $10.81 \pm 5.24$ | $15.56 \pm 11.61$ | $5.66 \pm 5.79$ |
| Jeong & Lim (2021)<sup>24</sup> | 0.7780 | $5.49 \pm 5.09$ | $6.63 \pm 6.74$ | $3.90 \pm 5.75$ |
| Joung et al. (2024)<sup>25</sup> | 0.3451 | $11.83 \pm 6.54$ | $14.61 \pm 12.20$ | $7.65 \pm 6.43$ |

Performance was benchmarked against an upper-bound model trained on ground-truth (bedside monitor) ECG and PPG inputs rather than wearable sensor signals. This ground-truth model attained an R² of 0.824 and an MAE of 4.81 ± 4.53 mmHg, with SBP and DBP MAEs of 5.76 ± 6.02 and 3.55 ± 4.98 mmHg, respectively. The relatively narrow gap between the sensor-based CNN/LSTM hybrid (*R*^2^ = 0.812, *MAE* = 4.94 mmHg) and the ground-truth input model (*R*^2^ = 0.824, *MAE* = 4.81 mmHg) suggests that the wearable sensor signals retain most of the hemodynamic information present in clinical-grade recordings, with only modest degradation attributable to sensor noise and temporal misalignment. Our evaluation of the CNN/LSTM hybrid model that includes static SBP/DBP covariates from the beginning of each 20-minute segment further improves model performance (*R*^2^ = 0.816, *MAE* = 4.90 mmHg; **Table S1**). We further observe a low degree of error in the distribution of ABP errors, as exhibited in **Figure S3**. We conduct a Bland-Altman agreement analysis for SBP and DBP, as shown in **Figure S4**, demonstrating our model shows a low degree of bias. Our model outperformed comparable baselines from Yen et al. (2022)^23^, Jeong & Lim (2021)^24^, and Joung et al. (2024)^25^ (see **Supplemental Methods**).

### Conformal prediction and uncertainty quantification

To further quantify the reliability of waveform predictions, we applied split-conformal prediction to generate uncertainty intervals with formal coverage guarantees. Across the test set, the resulting intervals achieved empirical coverage closely matching the nominal target, demonstrating that the conformal framework was well-calibrated in this setting (**Figure 2**). Importantly, the interval widths varied systematically with the predicted blood pressure values, reflecting the use of a conditionally adaptive scaling function derived from calibration residuals. This led to wider intervals in regions of higher model uncertainty, particularly near systolic peaks, and narrower intervals during diastolic phases, providing a physiologically intuitive representation of uncertainty over the cardiac cycle. We demonstrate that the conformal prediction model demonstrates empirical coverage over the theoretically-specified (see **Figure S5**).

**Figure 2.**
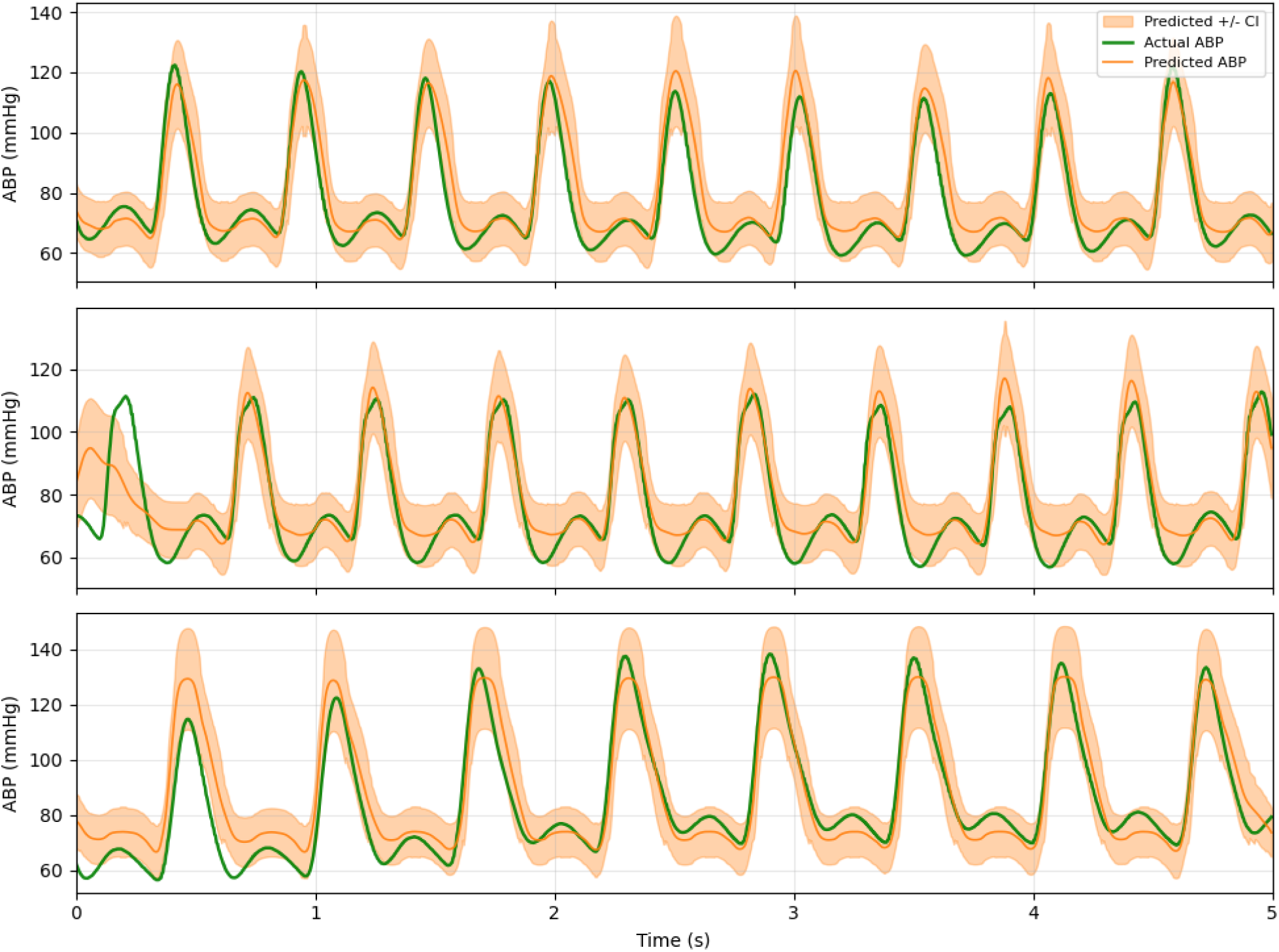
Conformal prediction intervals and empirical coverage for predicted arterial blood pressure waveforms. Representative 5-second test segments showing ground-truth invasive ABP (green), predicted ABP from the CNN/LSTM hybrid model (orange), and the split-conformal prediction band (shaded orange region,). Interval width varies across the cardiac cycle, reflecting the conditionally varying scale). estimated from calibration residuals binned by predicted value — intervals are wider near systolic peaks where model uncertainty is greatest and narrower near diastolic troughs.

To characterize model performance across the full hemodynamic range encountered in this critically ill population, we stratified segment-level MAE by binned ground-truth SBP, DBP, and MAP values (**Figure S6**). For SBP, prediction errors were lowest in the normotensive range (approximately 100–130 mmHg). At the extremes of the SBP distribution, errors increased modestly. A similar pattern was observed for DBP, with the lowest errors in the 50–70 mmHg range and slightly elevated errors at values above 90 mmHg or below 50 mmHg. MAP errors were relatively uniform across the central range (65–95 mmHg), with modest increases at the tails of the distribution.

To further characterize agreement between predicted and ground-truth blood pressure values, **Figure 3** shows scatter plots of segment-level systolic (SBP), diastolic (DBP), and mean arterial pressure (MAP) for the CNN/LSTM hybrid + BP model. Across all three measures, predictions are closely aligned with the identity line, indicating strong concordance over the full physiological range encountered in this cohort. The dispersion of points around the diagonal is modest and approximately symmetric, suggesting minimal systematic bias and consistent performance across low, normal, and high pressure values. This pattern is reflected in the high coefficients of determination and low mean absolute errors observed for each metric, with particularly tight agreement for MAP and DBP. Together, these results demonstrate that the model captures both the central tendency and variability of arterial pressure with high fidelity at the segment level.

**Figure 3.**
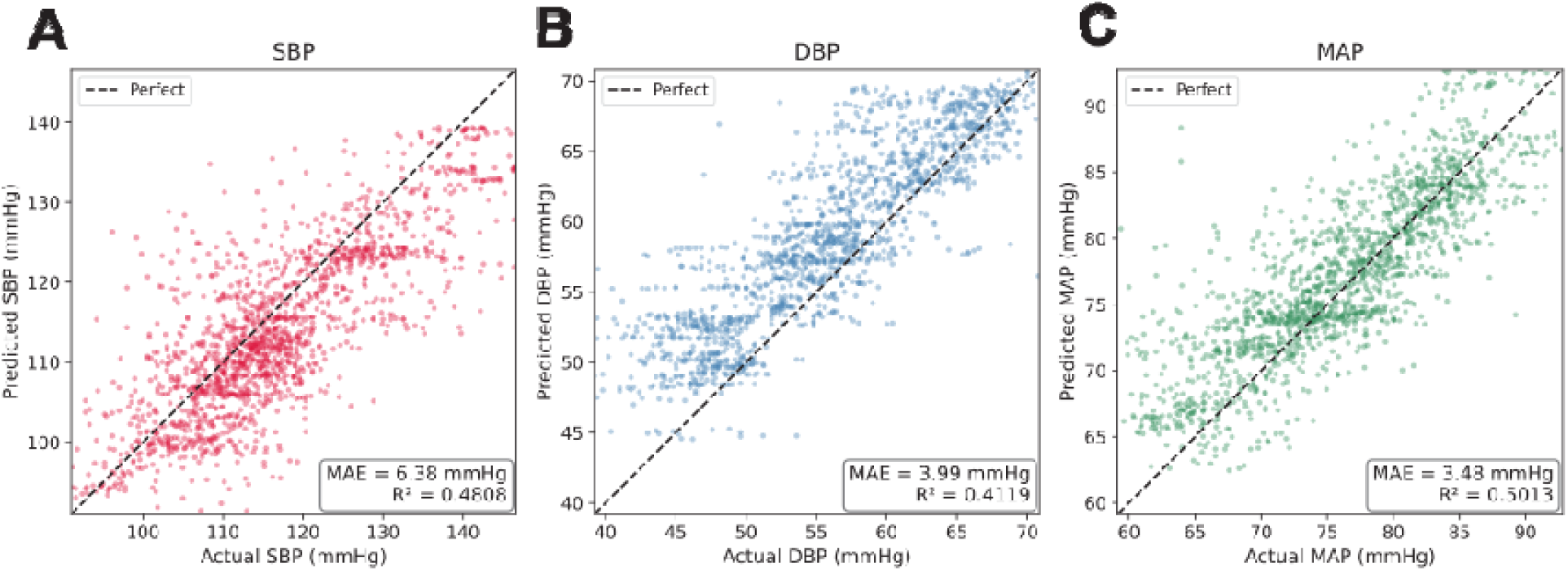
Mean absolute error of predicted blood pressure stratified by ground-truth blood pressure value. Segment-level mean absolute error (MAE, mmHg) for systolic blood pressure (SBP; A), diastolic blood pressure (DBP; B), and mean arterial pressure (MAP; C) as a function of the corresponding ground-truth value, for the CNN/LSTM hybrid + BP model. Each point represents the mean MAE within a binned range of ground-truth values. For visual concision, the median 99% of values are plotted.

## Discussion

In this study, we demonstrate that continuous arterial blood pressure waveforms in critically ill patients can be reconstructed with high fidelity from non-invasive PPG and ECG signals acquired by a wearable sensor array. The primary CNN/LSTM hybrid model achieved an *R*^2^ of 0.812 and a sample-level MAE of 4.94 mmHg, approaching the upper-bound performance obtained when clinical-grade bedside signals were used as inputs (*R*^2^ = 0.824, *MAE* = 4.81 mmHg). Beyond waveform-level accuracy, the split-conformal prediction framework provided calibrated, value-dependent uncertainty intervals that met or exceeded nominal coverage targets. Taken together, these findings suggest that wearable sensor-derived biosignals retain sufficient hemodynamic information to support continuous, non-invasive blood pressure estimation in a population characterized by the complex and rapidly changing physiology of critical illness.

Prior work in this domain has pursued two complementary research directions. The first focuses on predicting discrete blood pressure values: systolic (SBP), diastolic (DBP), and mean arterial pressure (MAP), from PPG and ECG signals using either model-driven approaches based on pulse wave velocity theory, or data-driven machine learning methods^26–29^. While these approaches have achieved promising accuracy in controlled settings, they provide only intermittent estimates and sacrifice the continuous waveform information available from invasive monitoring. The second research direction addresses this limitation by attempting to reconstruct the complete ABP waveform from non-invasive signals. Deep learning architectures including U-Net variants, variational autoencoders, and generative adversarial networks have demonstrated the feasibility of translating PPG signals to ABP waveforms, achieving mean absolute errors in the range of 2-5 mmHg for derived pressure values^30–33^. However, most existing studies have been conducted on databases of predominantly healthy subjects or general ICU populations without rigorous signal quality filtering, limiting their applicability to the complex hemodynamic states encountered in critically ill patients.

The systematic comparison across model architectures revealed several noteworthy patterns. Performance scaled with model capacity within the LSTM family, with the large LSTM (*R*^2^ = 0.787) outperforming the medium (*R*^2^ = 0.778) and small (*R*^2^ = 0.741) variants, consistent with the high dimensionality and temporal complexity of the waveform reconstruction task. The CNN/LSTM hybrid architecture further improved upon the pure LSTM models, suggesting that the convolutional front-end extracts local morphological features, such as the rising edge of the PPG pulse or QRS complex shape, that complement the temporal dynamics captured by the recurrent layers. By contrast, the U-Net architecture, which has been widely adopted in prior PPG-to-ABP translation studies, performed substantially worse in this cohort (*R*^2^ = 0.507). This discrepancy may reflect the fact that U-Net architectures were originally developed and validated on healthier populations or on databases with less hemodynamic variability; in the heterogeneous ICU setting studied here, the fully convolutional encoder–decoder may lack the capacity to model the long-range temporal dependencies and beat-to-beat variability that recurrent architectures handle more naturally.

The application of split-conformal prediction to ABP waveform estimation represents, to our knowledge, the first use of distribution-free uncertainty quantification in this domain. Existing approaches to PPG-based blood pressure estimation typically report only point estimates and aggregate error statistics, leaving clinicians without a principled way to assess the reliability of any individual prediction. The conformal framework addresses this gap by providing prediction intervals with finite-sample coverage guarantees under the assumption of exchangeability, a weaker assumption than the distributional assumptions required by Bayesian or parametric approaches. The conditionally varying scale further enhances interpretability: intervals are wider near systolic peaks, where calibration residuals tend to be largest, and narrower near diastolic troughs, providing a physiologically intuitive uncertainty profile that tracks the cardiac cycle. The tertile-based classification of interval width into narrow, moderate, and wide categories offers a practical mechanism for clinical decision support, enabling clinicians to distinguish predictions in which the model is relatively confident from those that should be interpreted with greater caution. In a clinical deployment, segments with wide uncertainty intervals could trigger confirmatory cuff measurements or flag the need for closer clinical attention, creating a hybrid monitoring paradigm that leverages the continuous nature of the wearable estimates while acknowledging their limitations.

Several limitations of this study should be acknowledged. First, the analysis cohort of 28 patients, while clinically diverse, is relatively small, and the generalizability of the models to larger and more heterogeneous populations remains to be established. The data were collected at a single academic medical center, and ICU practices, patient demographics, and hemodynamic profiles may differ across institutions. Second, the signal quality filters excluded a substantial proportion of the recorded data, and the model’s performance on lower-quality segments, which may be more representative of real-world ambulatory conditions, is not characterized. Third, recording durations were relatively short (mean 46 minutes), and the stability of model performance over longer monitoring sessions, during which physiological drift, sensor displacement, and changes in patient position may occur, has not been assessed.

Future work should address these limitations through multi-center validation with larger cohorts, prospective evaluation using oscillometric calibration, and extended monitoring sessions that assess temporal stability. The incorporation of additional sensor modalities available in the MOSAIC architecture, including accelerometry and electrodermal activity, may improve robustness to motion artifact and provide complementary hemodynamic information. Transfer learning or domain adaptation strategies could be explored to adapt models trained in the ICU to ambulatory or home-monitoring settings, where the distribution of hemodynamic states and signal quality characteristics differs substantially. Finally, the clinical, operational and economic impact of continuous non-invasive ABP monitoring, including the frequency of invasive line placement, time to therapeutic intervention for blood pressure derangements, incidence of catheter-related complications, and cost should be evaluated propsectively.

In conclusion, we have demonstrated that a CNN/LSTM hybrid deep learning model can reconstruct continuous arterial blood pressure waveforms from wearable PPG and ECG signals with accuracy approaching that of clinical-grade inputs. The model reliably detects clinically significant hypertensive and hypotensive episodes, and the conformal prediction framework provides calibrated uncertainty estimates that could support bedside decision-making. These results establish a foundation for the development of non-invasive, wearable hemodynamic monitoring systems that could reduce dependence on invasive arterial catheterization, expand access to continuous blood pressure assessment beyond the ICU, and ultimately improve the safety and precision of hemodynamic management in both hospital and community settings.

## Data and code availability

Code will be released upon reasonable request to the corresponding author. MOSAIC sensor data is protected by Johns Hopkins University privacy restrictions, and sharing of these data will require a Data Use Agreement consented to by the University.

## Acknowledgements

This research was made possible by the Johns Hopkins Institute for Clinical and Translational Research (ICTR), which is funded in part by Grant Number 1UM1TR004926 from the National Center for Advancing Translational Sciences (NCATS), a component of the National Institutes of Health (NIH), and NIH Roadmap for Medical Research. Its contents are solely the responsibility of the authors and do not necessarily represent the official view of the Johns Hopkins ICTR, NCATS or NIH.

This material is based upon work supported by the National Science Foundation Graduate Research Fellowship under Grant No. DGE2139757, awarded to CH. Any opinion, findings, and conclusions or recommendations expressed in this material are those of the authors(s) and do not necessarily reflect the views of the National Science Foundation.

## Ethics statement

All participants provided informed consent, either individually or through their legally authorized representative. Research was approved under the auspices of approved JHU IRB protocol IRB00384821.

## Supplementary Methods

### Comparison models

To contextualize the performance of the primary CNN/LSTM hybrid architecture, we evaluated a series of comparison models spanning naive baselines, pure recurrent architectures at varying capacities, a transformer-based model, a U-Net encoder-decoder, and a previously published CNN approach. All comparison models were trained and evaluated using the identical preprocessing pipeline, data splits, input standardization, and evaluation metrics described in the main text. All models received the same two-channel sequential input (sensor ECG and PPG, 1,250 time steps at 250 Hz).

#### Dummy regressor

The dummy regressor served as a naive baseline that predicted the training-set mean ABP value at every time point for every test segment, without using any input signal information. This model establishes a lower bound on performance and provides a reference against which all learned models can be compared.

#### LSTM models

We evaluated three pure LSTM architectures of increasing capacity to characterize the relationship between model size and waveform reconstruction accuracy. All three variants consisted of four stacked bidirectional LSTM layers, differing only in the number of hidden units per layer and the dimensionality of the static covariate projection. The small LSTM used hidden sizes of 32, 16, 8, and 4 units across the four layers, with a 4-dimensional static branch projection, matching the recurrent component of the primary CNN/LSTM hybrid model but omitting the convolutional front-end. The medium LSTM doubled the hidden sizes to 64, 32, 16, and 8 units, with an 8-dimensional static projection. The large LSTM further increased capacity to 128, 64, 32, and 16 hidden units, with a 16-dimensional static projection. Dropout and recurrent dropout were set to 0.2 for the first three layers and 0.1 for the final layer across all three variants. Training hyperparameters, including the Adam optimizer (learning rate 0.001), early stopping (patience 5 epochs), learning rate reduction on plateau (factor 0.5, patience 3 epochs, minimum learning rate 1 × 10□□), batch size (64), and weighted mean squared error loss, were identical to those used for the primary model.

#### Transformer model

To evaluate whether attention-based sequence modeling could match or exceed recurrent architectures for this task, we implemented a transformer encoder model. The two-channel input sequence was first projected to a higher-dimensional embedding space via a linear layer, after which learned positional encodings were added to preserve temporal order. The embedded sequence was passed through a stack of transformer encoder layers, each comprising multi-head self-attention and a position-wise feed-forward network with GELU activation, layer normalization, and residual connections. A final linear projection layer mapped the transformer output to the predicted ABP waveform at each time step. When the static BP covariate branch was included, the block-level SBP and DBP values were projected through a dense layer with ReLU activation, broadcast along the time dimension, and concatenated with the transformer encoder output prior to the final projection, following the same fusion strategy used in the CNN/LSTM hybrid. Dropout was applied within the attention layers and feed-forward blocks.

#### U-Net model

The U-Net architecture was included as a representative fully convolutional encoder–decoder approach, motivated by its widespread adoption in prior PPG-to-ABP translation studies. The model followed the standard U-Net design with a contracting encoder path and an expansive decoder path connected by skip connections at each resolution level. The encoder consisted of repeated blocks of one-dimensional convolutions, batch normalization, ReLU activation, and max pooling, progressively reducing the temporal resolution while increasing the number of feature channels. The decoder mirrored this structure with transposed convolutions for upsampling, with skip connections concatenating encoder feature maps at corresponding resolution levels to preserve fine-grained temporal detail. A final 1X 1 convolutional layer projected the decoder output to a single-channel predicted ABP waveform matching the input length (1,250 samples). For the variant incorporating static BP covariates, block-level SBP and DBP values were projected through a dense layer, broadcast along the time dimension, and concatenated with the bottleneck representation between the encoder and decoder paths. Training hyperparameters were consistent with all other models.

#### Ground-truth input models

To establish an upper bound on achievable performance — representing the best-case scenario in which sensor noise, signal degradation, and temporal misalignment are absent — we trained the CNN/LSTM hybrid architecture using ground-truth ECG and PPG signals from the bedside monitor as inputs, rather than signals from the MOSAIC wearable sensors. All other aspects of the model, including architecture, hyperparameters, preprocessing, and evaluation, were identical to the primary sensor-input model. This ground-truth model was evaluated both with and without block-level BP covariates (GT and GT + BP, respectively). The performance gap between the ground-truth input model and the corresponding sensor-input model quantifies the information loss attributable to the wearable sensing modality, including factors such as reduced signal-to-noise ratio, motion artifact, and residual temporal misalignment after the R-peak–based registration procedure. A narrow gap indicates that the MOSAIC sensors capture most of the hemodynamically relevant information present in clinical-grade recordings.

### Comparison models

To benchmark against a previously published approach, we re-implemented architectures as described in the methods sections of Yen, Chang and Liao ^23^, Joung, Jung, Lee, Chae, Kim, Park, Shin, Kim, Lee and Choi ^25^, and Jeong and Lim ^24^. All models were trained using the same training and evaluation paradigm as our models. We note, however, that code was not publicly available for any of the three models – while we attempted to faithfully replicate the architectures described, some modifications were required to fit our paradigm.

## Supplementary Tables

**Table S1.** Results for the models including static blood pressure measurements. Analogous table to Table 2, for models including static SBP/DBP values derived from the invasive ABP monitor.

| Model | $R^2$ | MAE | SBP MAE | DBP MAE |
| --- | --- | --- | --- | --- |
| GT + BP | 0.835 | $4.72 \pm 4.32$ | $5.53 \pm 6.04$ | $3.33 \pm 5.02$ |
| LSTM (small) + BP | 0.759 | $6.01 \pm 5.09$ | $7.43 \pm 7.33$ | $3.82 \pm 5.35$ |
| LSTM (med) + BP | 0.774 | $5.67 \pm 5.04$ | $7.25 \pm 7.01$ | $3.93 \pm 5.56$ |
| LSTM (large) + BP | 0.799 | $5.12 \pm 5.05$ | <u>$6.25 \pm 7.01$</u> | <u>$3.12 \pm 5.24$</u> |
| Transformer + BP | 0.788 | $5.28 \pm 5.27$ | $7.24 \pm 6.82$ | **$3.06 \pm 5.38$** |
| CNN/LSTM hybrid + BP | **0.816** | **$4.90 \pm 4.87$** | **$6.12 \pm 6.55$** | $3.98 \pm 5.58$ |
| UNET + BP | 0.612 | $7.98 \pm 4.67$ | $9.07 \pm 7.91$ | $3.05 \pm 5.14$ |

## Supplementary Figures

**Figure S1.**
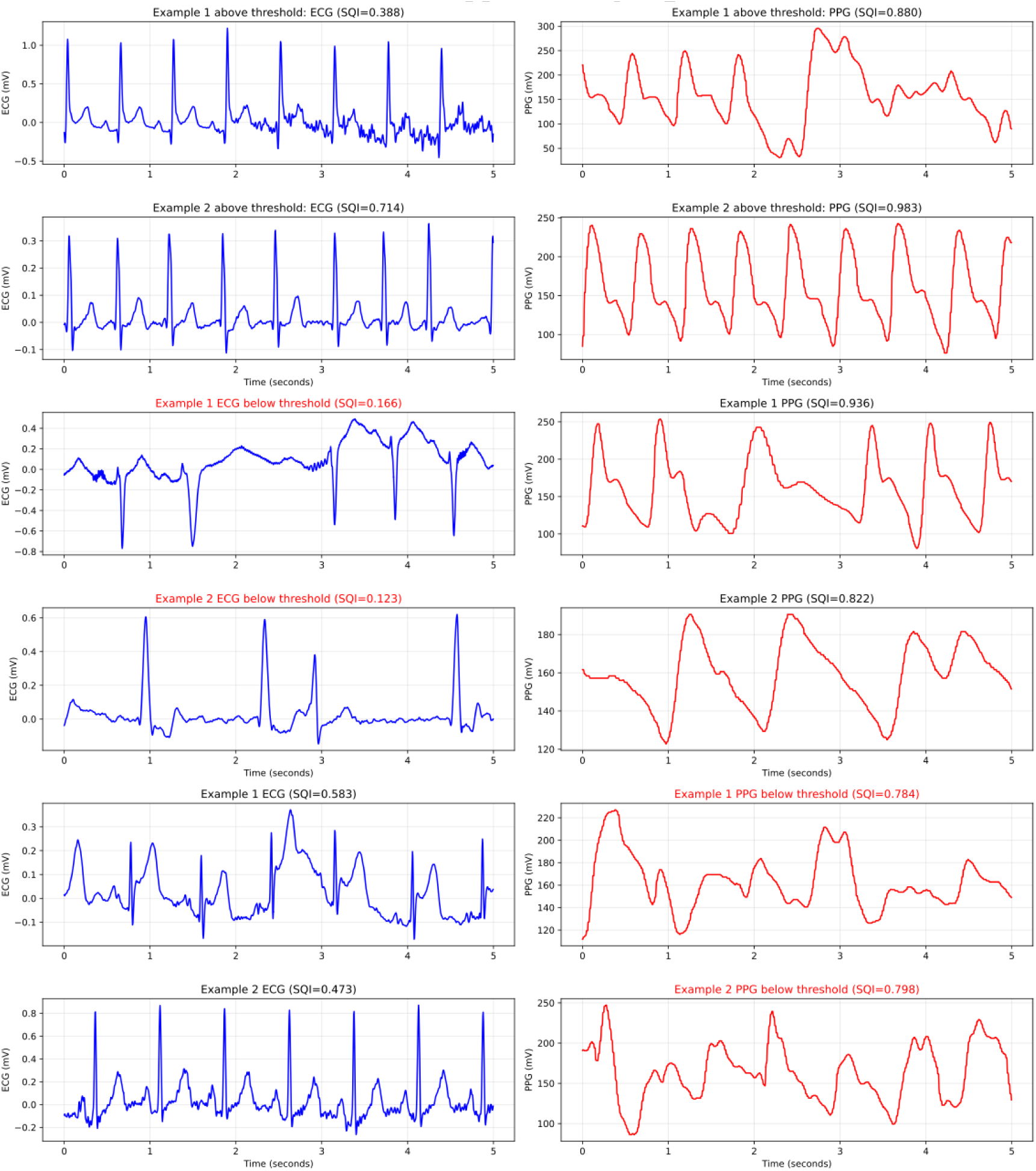
Signal quality assessment and impact of signal quality index (SQI) thresholds on sample retention and model performance. Distribution of per-segment ECG and PPG signal quality indices (SQI) across the study cohort. SQI was computed for ECG using the average-QRS quality metric and for PPG using the template-match quality metric, with the mean over each 5-second segment taken as the segment-level SQI. Segments were retained for analysis only if both ECG SQI and PPG SQI met or exceeded a threshold of 0.6. The figure illustrates the distribution of SQI values across all candidate segments and the proportion of segments excluded at various threshold levels, providing context for the trade-off between data quality and sample size in the final analysis cohort.

**Figure S2.**
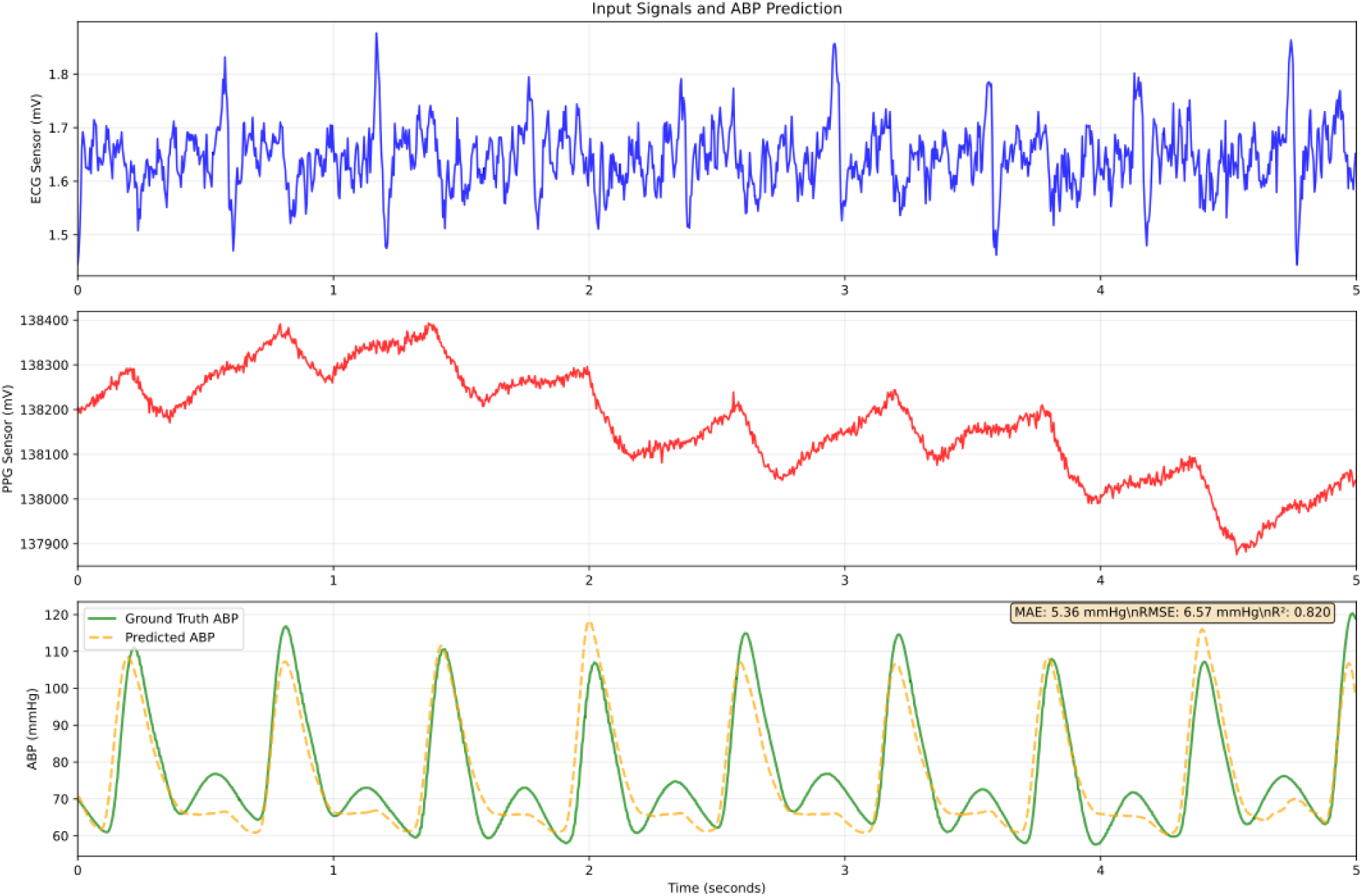
Example arterial blood pressure waveform predictions from the CNN/LSTM hybrid + BP model. Representative 5-second test segments showing the input wearable sensor signals (ECG and PPG), the ground-truth invasive arterial blood pressure (ABP) waveform from the bedside monitor, and the corresponding model-predicted ABP waveform. The close correspondence between predicted and ground-truth waveforms demonstrates the model’s ability to reconstruct beat-to-beat ABP dynamics, including systolic upstrokes, dicrotic notch features, and diastolic decay, from non-invasive biosignals.

**Figure S3.**
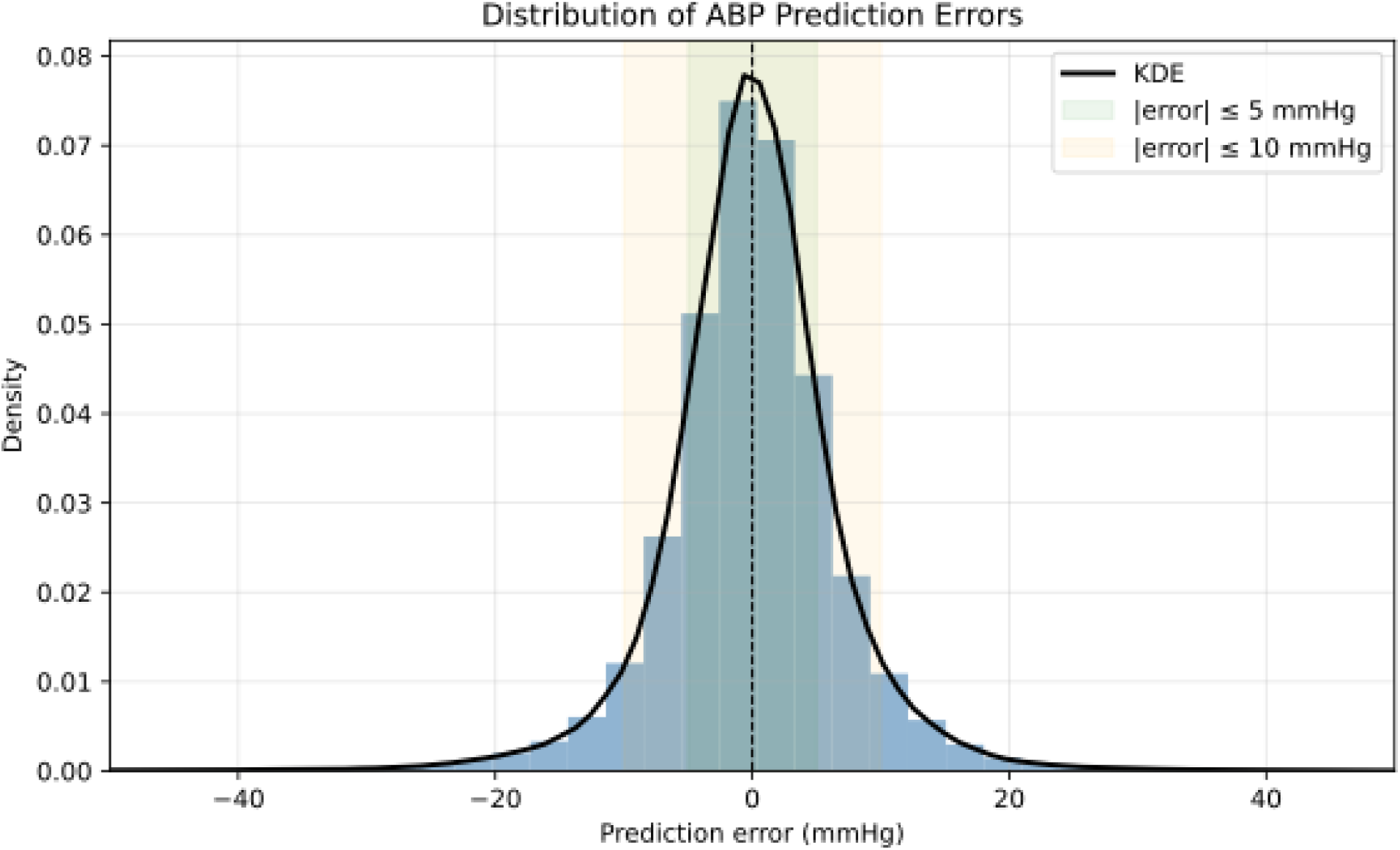
Distribution of arterial blood pressure prediction errors. Histogram of sample-level and segment-level prediction errors (predicted minus ground-truth ABP, in mmHg) for the CNN/LSTM hybrid + BP model on the held-out test set. Distributions are shown for overall waveform error as well as for derived systolic blood pressure (SBP) and diastolic blood pressure (DBP) errors.

**Figure S4.**
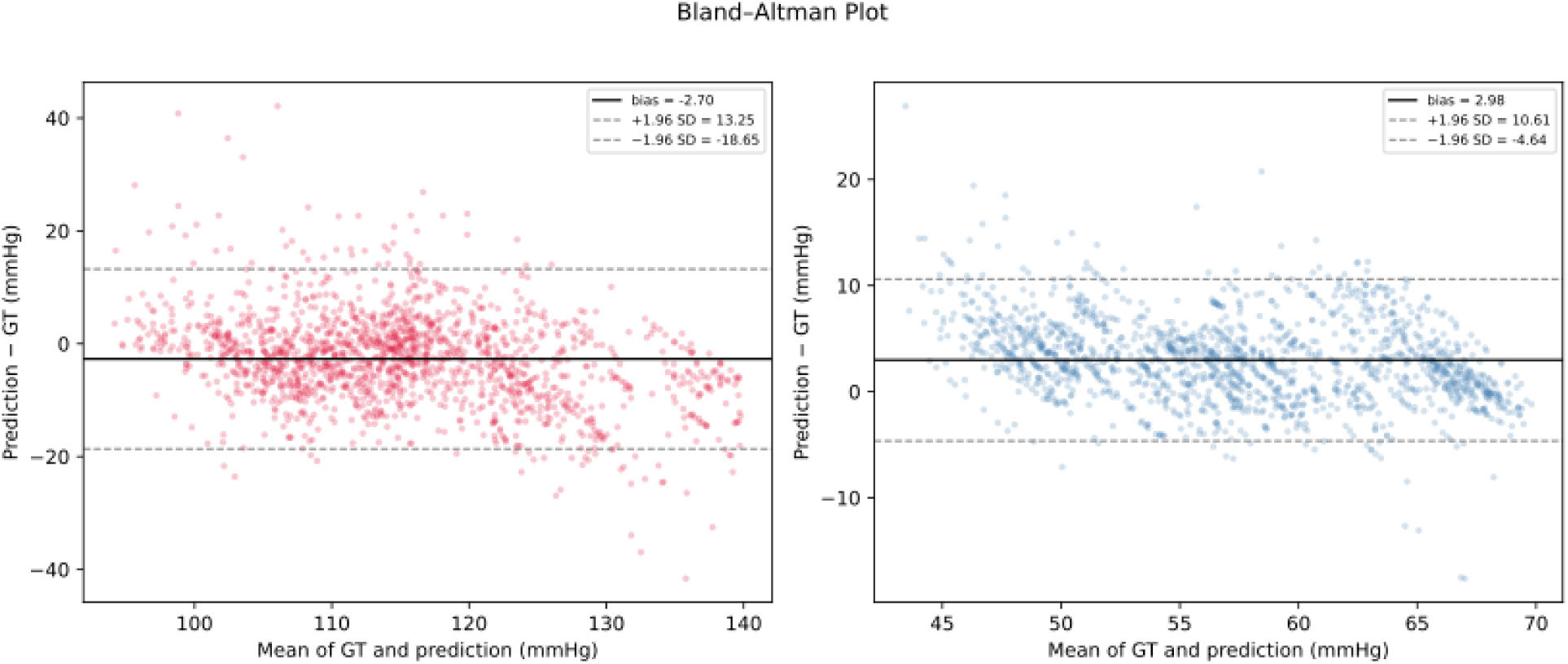
Bland-Altman agreement for segment-level systolic and diastolic blood pressure estimates. Bland-Altman plots comparing predicted versus ground-truth segment-level systolic blood pressure (SBP; left) and diastolic blood pressure (DBP; right) for the CNN/LSTM hybrid + BP model on the held-out test set. For visual simplicity, the median 99% of data points are plotted. Each point represents a 5-second segment, with the x-axis showing the mean of the predicted and ground-truth values and the y-axis showing their difference (prediction − ground truth, mmHg). Solid horizontal lines indicate the mean bias, and dashed lines indicate the 95% limits of agreement (bias ± 1.96 SD). The plots show overall agreement across the observed pressure range, with wider dispersion at the extremes.

**Figure S5.**
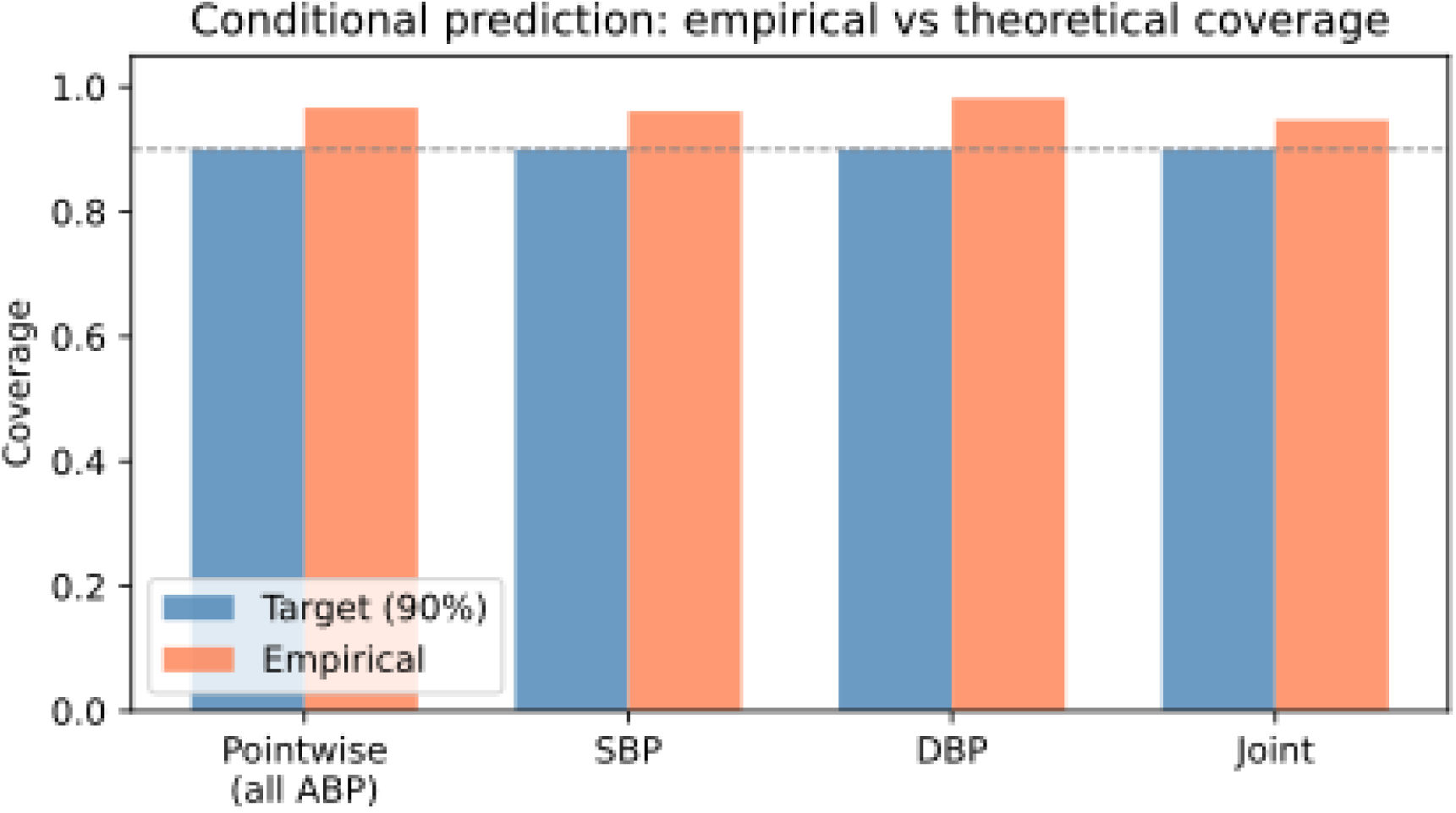
Conformal prediction coverage under patient-level data splitting. Comparison of nominal target coverage (90%, blue) and empirical coverage (orange) for the split-conformal prediction intervals generated by the CNN/LSTM hybrid + BP. Coverage is reported for pointwise ABP samples (all time points across all test segments), segment-level SBP, segment-level DBP, and joint wise (i.e., SBP + DBP) coverage. Pointwise empirical coverage closely matches the 90% target, while segment-level SBP and DBP coverage modestly exceeds the nominal level (∼92.5%), indicating that the conformal intervals remain well-calibrated or slightly conservative.

**Figure S6.**
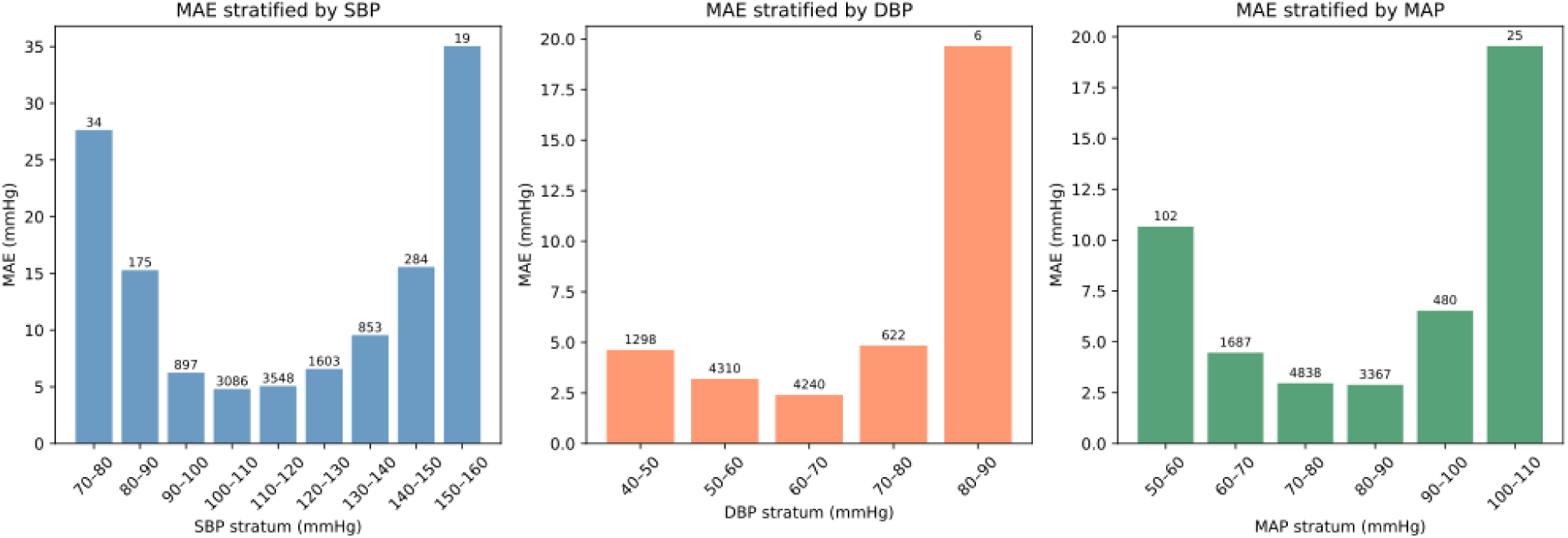
Mean prediction error stratified by ground-truth blood pressure value across model architectures. Segment-level mean absolute error (MAE, mmHg) for systolic blood pressure (SBP), diastolic blood pressure (DBP), and mean arterial pressure (MAP) as a function of the corresponding ground-truth value. Each point represents the mean MAE within a binned range of ground-truth values.

## References

1. Organization WH. Global report on hypertension: the race against a silent killer. World Health Organization; 2023.

2. Mills KT, Stefanescu A, He J. The global epidemiology of hypertension. Nature reviews nephrology. 2020;16(4):223–237.

3. Kirkland EB, Heincelman M, Bishu KG, et al. Trends in healthcare expenditures among US adults with hypertension: national estimates, 2003–2014. Journal of the American Heart Association. 2018;7(11):e008731.

4. Guerrero-García C, Rubio-Guerra AF. Combination therapy in the treatment of hypertension. Drugs in context. 2018;7:212531.

5. Bress AP, Anderson TS, Flack JM, et al. The management of elevated blood pressure in the acute care setting: a scientific statement from the American Heart Association. Hypertension. 2024;81(8):e94–e106.

6. Peixoto AJ. Acute severe hypertension. New England Journal of Medicine. 2019;381(19):1843–1852.

7. Saugel B, Dueck R, Wagner JY. Measurement of blood pressure. Best practice & research Clinical anaesthesiology. 2014;28(4):309–322.

8. Scheer BV, Perel A, Pfeiffer UJ. Clinical review: complications and risk factors of peripheral arterial catheters used for haemodynamic monitoring in anaesthesia and intensive care medicine. Critical care. 2002;6(3):199.

9. Mukkamala R, Hahn J-O, Inan OT, et al. Toward ubiquitous blood pressure monitoring via pulse transit time: theory and practice. IEEE transactions on biomedical engineering. 2015;62(8):1879–1901.

10. Frezza EE, Mezghebe H. Indications and complications of arterial catheter use in surgical or medical intensive care units: analysis of 4932 patients. The American Surgeon. 1998;64(2):127.

11. Cousins TR, O’Donnell JM. Arterial cannulation: a critical review. AANA journal. 2004;72(4)

12. O’grady NP, Alexander M, Burns LA, et al. Guidelines for the prevention of intravascular catheter-related infections. Clinical infectious diseases. 2011;52(9):e162–e193.

13. Elgendi M, Fletcher R, Liang Y, et al. The use of photoplethysmography for assessing hypertension. NPJ digital medicine. 2019;2(1):60.

14. Block RC, Yavarimanesh M, Natarajan K, et al. Conventional pulse transit times as markers of blood pressure changes in humans. Scientific Reports. 2020;10(1):16373.

15. Escobar-Restrepo B, Torres-Villa R, Kyriacou PA. Evaluation of the linear relationship between pulse arrival time and blood pressure in ICU patients: Potential and limitations. Frontiers in physiology. 2018;9:1848.

16. Wesseling K. Finapres, continuous noninvasive finger arterial pressure based on the method of Peñáz. Blood Pressure Measurements: New Techniques in Automatic and 24-hour Indirect Monitoring. Springer; 1990:161–172.

17. McAuley D, Silke B, Farrell S. Reliability of blood pressure determination with the Finapres with altered physiological states or pharmacodynamic conditions. Clinical Autonomic Research. 1997;7(4):179–184.

18. Rattray J. Continuous Health Monitoring with Distributed, Networked Wearables. Johns Hopkins University; 2024.

19. Rattray J, Nnadi B, Rapuri S, et al. A Wearable Multi-modal Sensor Array for Continuous Cuffless Blood Pressure Estimation. medRxiv. 2026:2026.01. 25.26344788.

20. Makowski D, Pham T, Lau ZJ, et al. NeuroKit2: A Python toolbox for neurophysiological signal processing. Behavior research methods. 2021;53(4):1689–1696.

21. Pan J, Tompkins WJ. A real-time QRS detection algorithm. IEEE transactions on biomedical engineering. 2007;(3):230–236.

22. Elgendi M, Norton I, Brearley M, Abbott D, Schuurmans D. Systolic peak detection in acceleration photoplethysmograms measured from emergency responders in tropical conditions. PloS one. 2013;8(10):e76585.

23. Yen C-T, Chang S-N, Liao C-H. Estimation of beat-by-beat blood pressure and heart rate from ECG and PPG using a fine-tuned deep CNN model. IEEE Access. 2022;10:85459–85469.

24. Jeong DU, Lim KM. Combined deep CNN–LSTM network-based multitasking learning architecture for noninvasive continuous blood pressure estimation using difference in ECG-PPG features. Scientific Reports. 2021;11(1):13539.

25. Joung J, Jung C-W, Lee H-C, et al. Continuous cuffless blood pressure monitoring using photoplethysmography-based PPG2BP-net for high intrasubject blood pressure variations. Scientific reports. 2023;13(1):8605.

26. Kachuee M, Kiani MM, Mohammadzade H, Shabany M. Cuffless blood pressure estimation algorithms for continuous health-care monitoring. IEEE Transactions on Biomedical Engineering. 2016;64(4):859–869.

27. Slapničar G, Mlakar N, Luštrek M. Blood pressure estimation from photoplethysmogram using a spectro-temporal deep neural network. Sensors. 2019;19(15):3420.

28. El-Hajj C, Kyriacou PA. A review of machine learning techniques in photoplethysmography for the non-invasive cuff-less measurement of blood pressure. Biomedical Signal Processing and Control. 2020;58:101870.

29. Zhang Y, Feng Z. A SVM method for continuous blood pressure estimation from a PPG signal. 2017:128–132.

30. Ibtehaz N, Mahmud S, Chowdhury ME, et al. PPG2ABP: Translating photoplethysmogram (PPG) signals to arterial blood pressure (ABP) waveforms. Bioengineering. 2022;9(11):692.

31. Athaya T, Choi S. An estimation method of continuous non-invasive arterial blood pressure waveform using photoplethysmography: A U-Net architecture-based approach. Sensors. 2021;21(5):1867.

32. Cheng J, Xu Y, Song R, Liu Y, Li C, Chen X. Prediction of arterial blood pressure waveforms from photoplethysmogram signals via fully convolutional neural networks. Computers in Biology and Medicine. 2021;138:104877.

33. Qin K, Huang W, Zhang T. Deep generative model with domain adversarial training for predicting arterial blood pressure waveform from photoplethysmogram signal. Biomedical Signal Processing and Control. 2021;70:102972.

